# Symptoms persisting after hospitalization for COVID-19: 12 months interim results of the COFLOW study

**DOI:** 10.1101/2021.12.11.21267652

**Authors:** L.M. Bek, J.C. Berentschot, M.H. Heijenbrok-Kal, S. Huijts, M.E. van Genderen, J.H. Vlake, J. van Bommel, J.G.J.V. Aerts, G.M. Ribbers, H.J.G. van den Berg-Emons, M.E. Hellemons, on behalf of the CO-FLOW collaboration Group

**Author notes:** **Correspondence:** M.E. Hellemons, Department of Respiratory Medicine, Erasmus Medical Centre, Dr. Molewaterplein 40, 3015 GD Rotterdam, The Netherlands. Authors share first authorship (L.M. Bek, J.C. Berentschot). Share senior authorship (H.J.G. van den Berg-Emons, M.E. Hellemons). Collaboration Group authors mentioned on last page.

## Abstract

**Introduction:** A large proportion of patients experiences a wide range of sequelae after acute COVID-19 infection, especially after severe illness. The long-term health sequelae need to be assessed. Our objective was to longitudinally assess persistence of symptoms and clusters of symptoms up to 12 months after hospitalization for COVID-19, and to assess determinants of the main persistent symptoms.

**Methods:** In this multicenter prospective cohort study patients with COVID-19 are followed up for 2 years with measurements at 3, 6, 12, and 24 months after hospital discharge. Here, we present interim results regarding persistent symptoms up to 12 months. Symptoms were clustered into physical, respiratory, cognitive and fatigue symptoms.

**Results:** We included 492 patients; mean age was 60.2±10.7 years, 335 (68.1%) males, median length of hospital stay 11 (6.0-27.0) days. At 3 months after discharge 97.0% of the patients had at least 1 persisting symptom, this declined to 95.5% and 92.0% at 6 and 12 months, respectively (p=0.010). Muscle weakness, exertional dyspnea, fatigue, and memory and concentration problems were the most prevalent symptoms with rates over 50% during follow-up. Over time, muscle weakness, hair loss, and exertional dyspnea decreased significantly (p<0.001), while other symptoms, such as fatigue, concentration and memory problems, anosmia, and ageusia persisted. Symptoms from the physical and respiratory cluster declined significantly over time, in contrast to symptoms from the fatigue and cognitive clusters. Female gender was the most important predictor of persistent symptoms and co-occurrence of symptoms from all clusters. Shorter hospital stay and treatment with steroids were related with decreased muscle weakness; comorbidity and being employed were related with increased fatigue; and shorter hospital stay and comorbidity were related with memory problems.

**Conclusion:** The majority of patients experienced COVID-19 sequelae up to 12 months after hospitalization. Whereas physical and respiratory symptoms showed slow gradual decline, fatigue and cognitive symptoms did not evidently resolve over time. This finding stresses the importance of finding the underlying causes and effective treatments for post-COVID condition, beside adequate COVID-19 prevention.

## INTRODUCTION

Acute coronavirus disease 2019 (COVID-19) infection in humans is associated with a heterogeneous range of symptoms including respiratory, musculoskeletal, gastrointestinal, and neurological symptoms. In 5-14% of patients the respiratory consequences of COVID-19 are severe, requiring hospitalization for oxygen supplementation or even prolonged ventilatory support (1).

Whereas a proportion of patients fully recovers, it becomes increasingly clear that a proportion of patients experiences a wide range of long-lasting sequelae after acute COVID-19 infection. Different terms are currently used for describing the presence of post-COVID-19 symptoms, such as long COVID, long haulers, post-COVID-19 syndrome, persistent post-COVID, and post-acute sequelae of COVID (PASC). Although several definitions are in place, persistent symptoms after COVID-19 are regarded as post-COVID-19 syndrome if they persist or present within 12⍰weeks of the onset of acute COVID-19 and last for at least 2 months, and are not attributable to alternative diagnoses (2, 3). The more recent World Health Organization (WHO) definition of post COVID-19 condition (PCC) is very similar to this definition, adding that symptoms may be new onset following initial recovery from an acute COVID-19 episode or persist from the initial illness and must persist for at least two months (4). Symptoms may also fluctuate or relapse over time. These post-acute COVID-19 sequelae encompass a wide range of symptoms and organs systems. Common symptoms include fatigue, shortness of breath and cognitive dysfunction (3, 4).

Although exact overall prevalence of these long-term symptoms remains unclear, it is estimated that between 2.6% and 13.7% of symptomatic patients experience persistent symptoms beyond 12 weeks after COVID-19 infection (5, 6). This number increases when patients are more severely affected (7). A recent systematic review described that more than 50% of all patients (the majority after hospitalization) experience post-acute COVID-19 sequelae, even up to 6 months after acute infection (8). Recent data from a Chinese cohort demonstrated that symptoms were still present in over 68% of hospitalized patients at 6 months after disease onset, decreasing to 49% at 12 months (9, 10).

The nature of the reported symptoms is diverse and ranges from exertional dyspnea to sensory overload. Although studies have tried to phenotype the patients with residual symptoms, looked into co-occurrence of pairs of post-COVID-19 symptoms, or report on assays of symptoms according to various organ systems, it remains unclear how the various domains of symptoms relate to each other and how frequently certain types of symptoms overlap (11-13).

Currently, most reports on persistent symptoms remain limited to 6 months after infection and little is known regarding the determinants of persistent symptoms. Also, most studies are cross-sectional and studies reporting outcomes across multiple time-points are scarce.

The aim of the current study was therefore to assess persistence of symptoms up to 12 months after discharge, to evaluate how persistent symptoms and clusters of symptoms develop over time, to determine how various clusters of symptoms overlap with each other, and to assess determinants of the main persistent symptoms after COVID-19.

## METHODS

### Study design

The COvid-19 Follow-up care paths and Long-term Outcomes Within the Dutch health care system (CO-FLOW) study is an ongoing multicenter prospective cohort study following COVID-19 patients discharged from hospitals in the Rotterdam-Rijnmond-Delft region in the Netherlands. Detailed description of its protocol can be found elsewere (14). In short, up to 2 years after hospitalization patients with COVID-19 are evaluated at 3, 6, 12, and 24 months after hospital discharge. Here, we present interim results regarding persisting symptoms obtained in the period from July 1^st^ 2020 until December 1^st^ 2021 as part of the CO-FLOW study up to 12 months after discharge. The Medical Ethics Committee of the Erasmus Medical Center (MC) approved this study (MEC-2020-0487). The trial was registered at The Netherlands Trial Register (NL8710), https://www.trialregister.nl, on June 12, 2020.

Adult patients (≥18 years of age) were eligible to participate in the CO-FLOW study if they had been hospitalized for COVID-19 (diagnosis based on either positive reverse transcription polymerase chain reaction or a clinical diagnosis combined with positive serology for COVID-19) within the previous 6 months and patient or relative had sufficient knowledge of the Dutch or English language. Incapacitated patients were unable to participate given the study procedures. For this study only participants with at least two study visits were included.

### Study Procedures

Patients were recruited during outpatient follow-up after discharge in one of the participating centers, at the inpatient rehabilitation center, or at the skilled nursing facility. All patients provided written informed consent before the start of the measurements. Study visits were synchronized with the patient’s regular follow-up for COVID-19 at each of the participating centers if possible. When patients were discharged from regular follow-up, study visits continued in the Erasmus University MC or, if patients were unable to come to the Erasmus University MC, a research assistant performed the study visit at home. During study visits patients performed non-invasive clinical tests, including physical, psychological, and cognitive evaluation. At 3, 6, and 12 months follow-up visits, patients received questionnaires via email or postal mail. Data were stored in Castor EDC (Castor EDC, Amsterdam, The Netherlands).

### Outcomes

A new Corona Symptom Checklist was developed for this study on ‘novel or worsened symptoms since the onset of COVID-19’ during the first three months of the study, based on the first experiences with post-COVID-19 patients. All questions are answered with “yes” or “no” (see appendix for complete questionnaire). During the study visits the Corona Symptom Checklist was administered by a research assistant in a face-to-face interview. As the Checklist was still under development when the study started amid the beginning of the COVID-19 pandemic, it was introduced at all study visits after August 5^th^ 2020. As the pandemic evolved and knowledge increased regarding PCC, additional questions were added (sensory overload, headache, chest pain) from June 2021 onwards.

As fatigue is considered as one of the most prevalent symptoms in PCC, we chose to report fatigue not based on the checklist results, but on the validated Fatigue Assessment Scale (FAS) that was assessed in all patients since study onset. The FAS is a 10-item self-report questionnaire and is validated in patients with chronic lung disease (15). The items are scored on a Likert scale ranging 1-5. A total score of ≥22 is considered to represent substantial fatigue and was used to indicate persisting fatigue (15).

Patient and clinical characteristics were collected at study visits and through electronic patient records. Patients characteristics included age, sex, body mass index (BMI), migration background, pre-COVID educational and employment status, smoking status, and comorbidities. Clinical characteristics included baseline laboratory and radiological parameters, complications during hospitalization including delirium and thrombosis, type and quantity of oxygen support, intensive care unit (ICU) admission, length of stay (LOS) ICU, LOS hospital, and COVID-19 directed treatment during hospital admission.

### Statistical analyses

We examined descriptive statistics to ensure data met statistical assumptions. Variables were presented as mean with standard deviation (SD), median with interquartile range, or numbers (n) with percentages (%) as appropriate.

Patient reported symptoms were clustered into one of four clusters according to the nature of the symptom: physical, respiratory, fatigue, and cognitive symptom cluster. The physical symptom cluster was composed of the symptoms muscle weakness, balance problems/dizziness, joint pain, tingling/numbness in extremities, hair loss, headache, chest pain, skin rash, vision problems, hoarseness, anosmia, ageusia, stool problems, claudication, hearing problems, and miction problems. The respiratory symptom cluster was composed of the symptoms exertional dyspnea, dyspnea, cough, and phlegm. The fatigue symptom clusters was composed of fatigue and sleeping problems. The cognitive symptom cluster was composed of the symptoms memory problems, concentration problems, sensory overload, and anxiety/nightmares. If any of the symptoms in the clusters was present at a time point, persisting symptoms in that cluster were scored as present at that time point. We used generalized estimating equations (GEE) with an unstructured covariance matrix to assess persistence of symptoms and symptom clusters over time. GEE accounts for correlations between patient follow-up measurements and includes all observed outcomes despite incomplete data. For longitudinal analyses, a Bonferroni correction was applied and a p-value <0.002 was considered statistically significant. Difference in the distribution of symptom clusters across gender were assessed with a Chi-Square test. Lastly, we performed multivariable logistic regression analyses with a backward selection procedure to determine which variables are independently associated with the most prevalent symptom per cluster at 3 months after discharge. The dependent variables were muscle weakness, deconditioning/exertional dyspnea, fatigue, and memory problems. We only reported determinants of symptoms at 3 months after discharge, as symptoms were most prevalent at 3 months after discharge and the majority did not decrease significantly over time. Determinants that were examined were age, sex, BMI, migration background (European, Dutch Caribbean, Asian, Turkish, and (North) African), pre-COVID educational (low, middle, high) and employment status (employed, not employed, retired), presence of comorbidity, smoking (never versus ever), BMI and C-reactive protein (CRP) at admission, complications of thrombosis or delirium, oxygen supplementation (none, nasal cannula or mask oxygen supplementation, high flow nasal cannula, mechanical ventilation), LOS hospital, and COVID-19 directed treatment with steroids. The covariates BMI and CRP were imputed with their mean value if missing. Variable elimination from the multivariable models was based on goodness of fit using the likelihood ratio test with a p-value of 0.1, and the final models are presented with adjusted odds ratios (OR) and 95% confidence interval (95%CI). All analyses were performed using Statistical Package for Social Sciences (SPSS) version 25 (IBM SPSS statistics, SPSS Inc, Chicago, IL, USA) and STATA version 8SE (StataCorp LLC, Texas, USA) and R version 4.1.1 (R-Foundation) were used for graphs.

## RESULTS

### Characteristics

Between July 1^st^ 2020 and December 1^st^ 2021, a total of 650 patients were included in COFLOW, of whom 492 participants underwent at least two study measurements and were included in this interim analysis. Baseline characteristics are presented in table 1. Patients had a mean age of 60.2±10.7 years, 335 (68.1%) were male, 403 (81.9%) had one or more comorbidities: most commonly obesity, cardiovascular, or pulmonary disease. Oxygen supplementation during admission was required by 474 (96.3%) patients, 199 (40.4%) had been admitted to the ICU, with a median LOS in ICU of 17 (9.0-30.5) days, and the median total LOS in the hospital was 11 (6.0-27.0) days. Of all patients, 357 (72.6%) received any COVID-19 directed treatment, of whom 330 (70.8%) received any form of steroids and 54 (11.5%) received directed anti-inflammatory treatment.

**Table 1.**
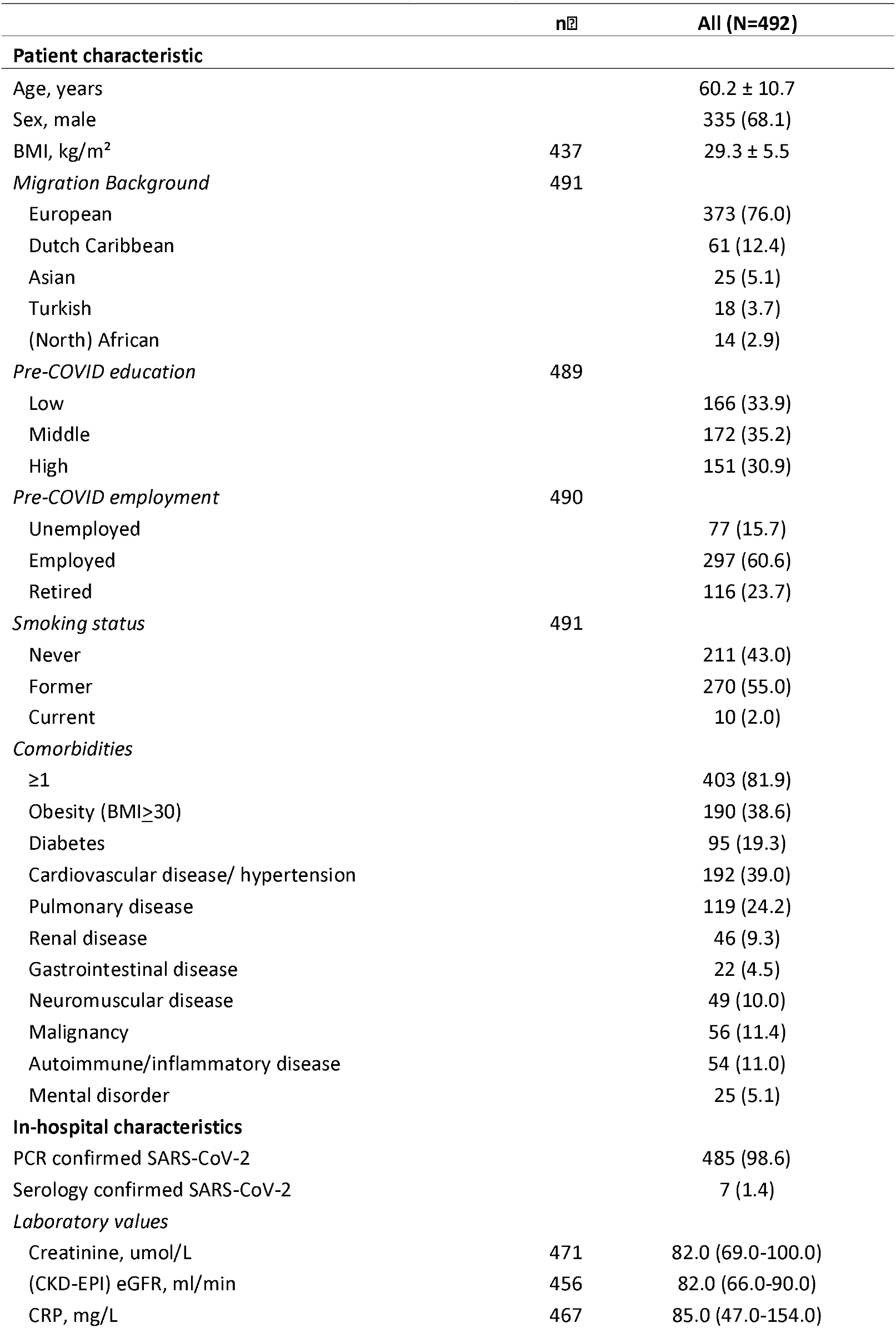
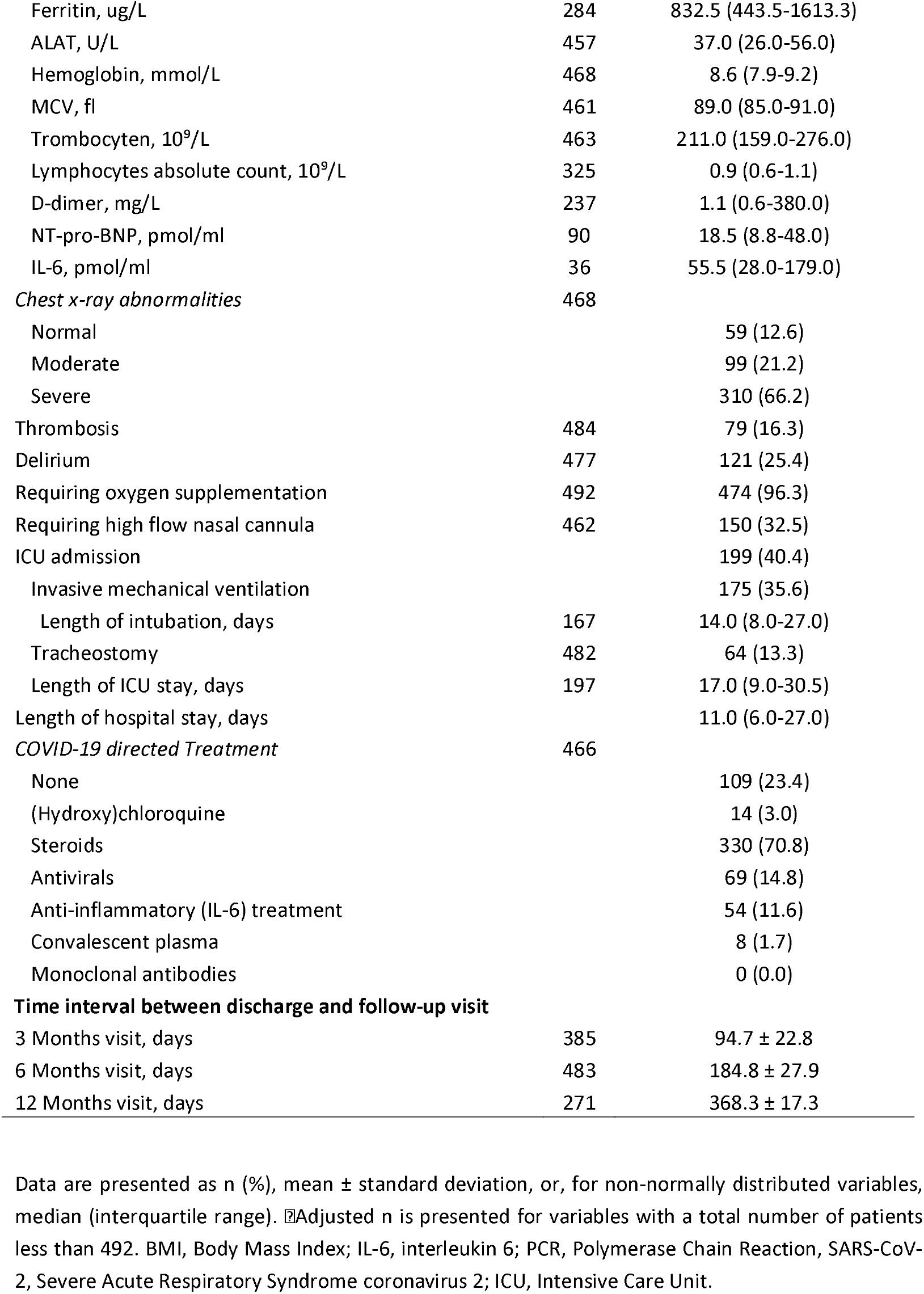
Patient and clinical characteristics of patients hospitalized for COVID-19

In total, 492 participant underwent at least two study measurements with the symptoms checklist. The flowchart of the included patients and study measurements is shown in figure 1.

**Figure 1:**
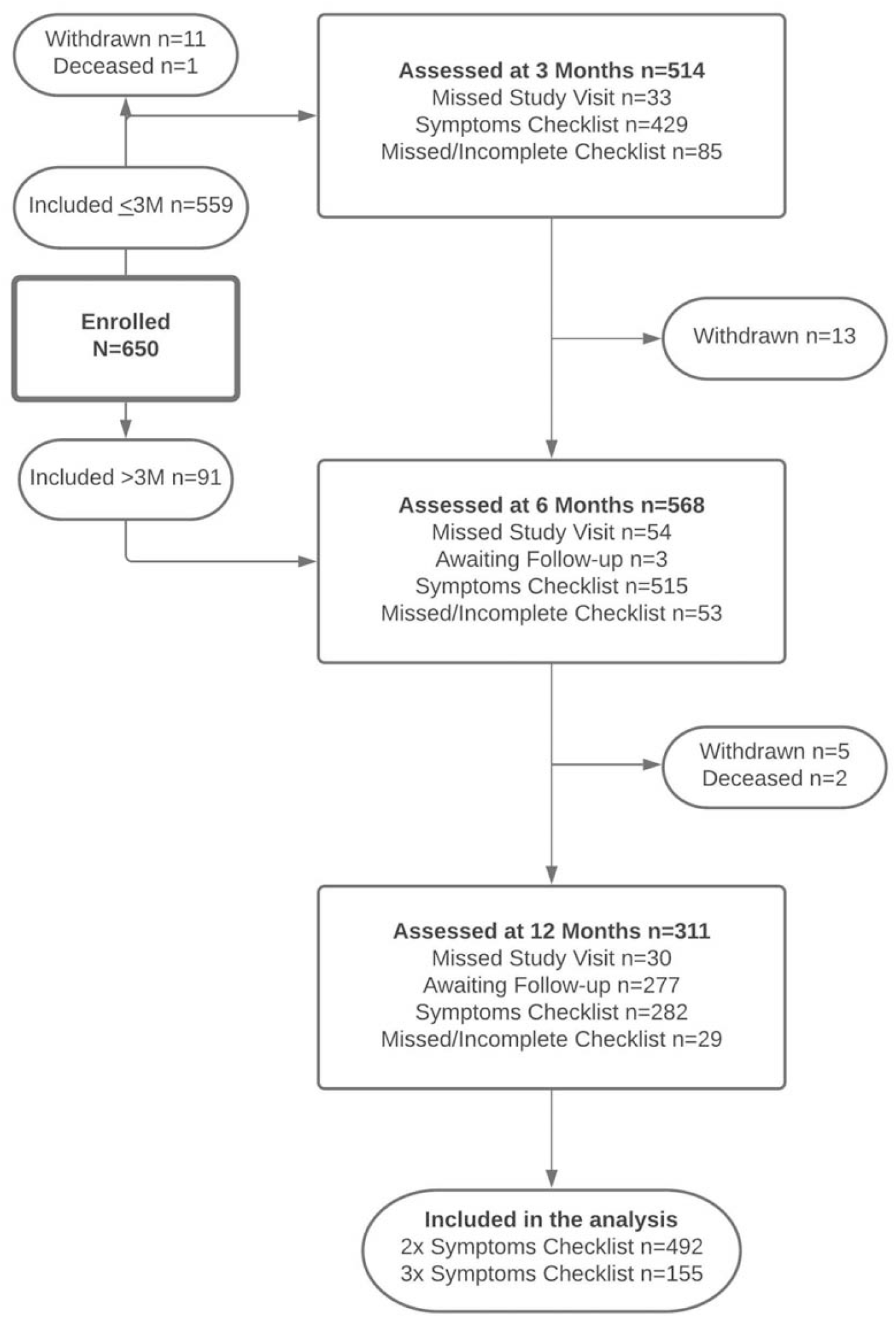
Flowchart of the patients in the CO-FLOW study during the interim analysis

### Persisting symptoms

Table 2 presents the number and proportion of patients with persisting symptoms at each follow-up measurement. At 3 months after discharge, 97.0% of the patients had at least 1 persisting symptom, this proportion of patients declined to 95.5% at 6 months and to 92.0% at 12 months (p=0.010). Presence of a single symptom varied from 9.7% for miction problems to 81.8% for exertional dyspnea. At all study visits, the most prevalent symptoms were muscle weakness, exertional dyspnea, fatigue, and memory and concentration problems. These symptoms were reported by more than 50% of the patients during follow-up; a large number of other persistent symptoms was frequently reported, presented in table 2 and figure 2. Symptoms that significantly declined over time were muscle weakness, hair loss, and exertional dyspnea (p<0.001).

**Table 2.** Prevalence of COVID-19 related symptoms at 3, 6, and 12 months follow-up in patients after hospitalization for COVID-19

|  | 3 Months<br>(n=385)<br>n (%) | 6 Months<br>(n=483)<br>n (%) | 12 Months<br>(n=271)<br>n (%) | p-value* |
| --- | --- | --- | --- | --- |
| **Physical symptoms** |  |  |  |  |
| Muscle weakness | 220 (57.1) | 234 (48.4) | 111 (41.0) | **<0.001** |
| Balance problems/dizziness | 169 (43.8) | 213 (44.4) | 116 (42.8) | 0.922 |
| Joint pain | 166 (43.2) | 201 (41.6) | 111 (41.0) | 0.352 |
| Tingling/numbness in extremities | 147 (36.8) | 163 (33.9) | 86 (32.1) | 0.291 |
| Hair loss | 138 (35.9) | 98 (20.3) | 35 (12.9) | **<0.001** |
| Headache* | 33 (31.4) | 57 (26.1) | 29 (18.6) | 0.579 |
| Chest pain* | 29 (29.0) | 40 (18.4) | 28 (17.8) | 0.069 |
| Skin rash | 99 (25.7) | 132 (27.4) | 82 (30.3) | 0.587 |
| Vision problems | 97 (25.2) | 148 (30.6) | 78 (28.8) | 0.023 |
| Hoarseness | 91 (23.6) | 125 (25.9) | 57 (21.0) | 0.088 |
| Anosmia | 84 (21.9) | 93 (19.3) | 53 (19.6) | 0.369 |
| Ageusia | 82 (21.2) | 94 (19.5) | 52 (19.2) | 0.185 |
| Stool problems | 68 (17.7) | 89 (18.5) | 41 (15.1) | 0.547 |
| Claudication | 54 (14.1) | 68 (14.1) | 27 (10.0) | 0.116 |
| Hearing problems | 52 (13.5) | 70 (14.5) | 53 (19.6) | 0.059 |
| Miction problems | 37 (9.7) | 58 (12.1) | 34 (12.5) | 0.269 |
| **Respiratory symptoms** |  |  |  |  |
| Exertional dyspnea | 315 (81.8) | 345 (71.4) | 171 (63.1) | **<0.001** |
| Dyspnea* | 78 (66.1) | 114 (51.8) | 83 (52.9) | 0.003 |
| Cough | 112 (29.0) | 119 (24.7) | 66 (24.4) | 0.329 |
| Phlegm | 98 (25.5) | 117 (24.2) | 67 (24.7) | 0.727 |
| **Fatigue symptoms** |  |  |  |  |
| Fatigue | 243 (64.5) | 277 (63.1) | 156 (60.2) | 0.932 |
| Sleeping problems | 141 (36.5) | 172 (35.6) | 96 (35.4) | 0.777 |
| **Cognitive symptoms** |  |  |  |  |
| Memory problems | 211 (54.7) | 271 (56.1) | 158 (58.3) | 0.144 |
| Concentration problems | 206 (53.4) | 249 (51.6) | 140 (51.7) | 0.826 |
| Sensory overload* | 44 (45.5) | 93 (43.9) | 58 (36.7) | 0.503 |
| Anxiety/nightmares | 56 (14.5) | 72 (14.9) | 40 (14.8) | 0.785 |

\*Symptoms headache, chest pain, dyspnea, and sensory overload were added in a later stage, resulting in lower total numbers. Data are presented as n (%) indicating the number of patients with symptoms. p-values are based on Generalized Estimating Equation analyses, with follow-up visit as fixed factor and symptom (yes/no) at each follow-up visit as dependent variable. Bonferroni correction was applied for multiple testing, a p-value < 0.002 was considered statistically significant (printed in bold).

**Figure 2:**
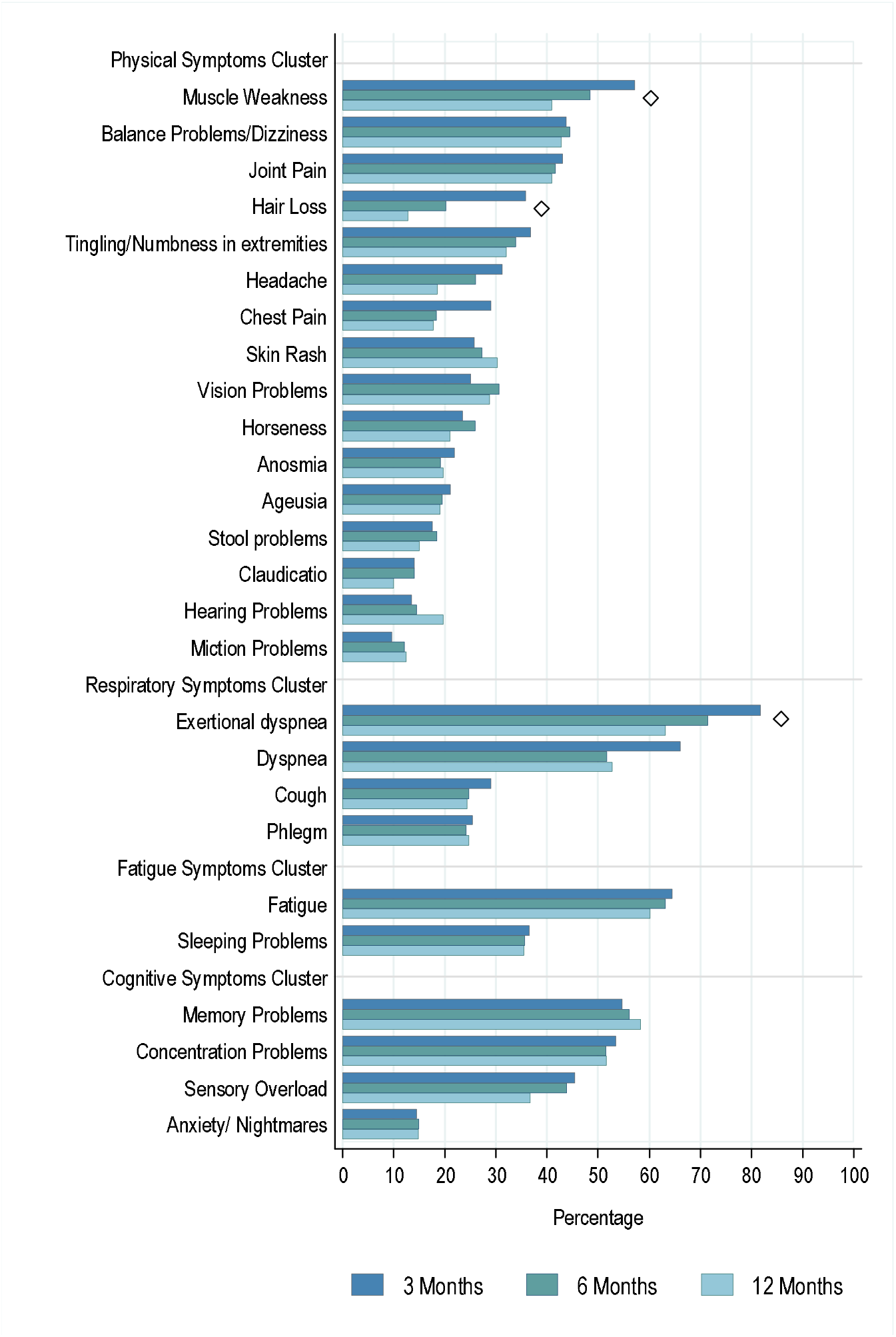
Symptom prevalence over time Prevalence of COVID-19-related symptoms at 3, 6, and 12 months follow-up in patients after hospitalization for COVID-19, sorted by symptoms cluster and from most to least frequently reported. Data are presented percentage of patients with symptoms. Symptoms marked with ♦ declined significantly over time based on Generalized Estimating Equation analyses, with follow-up visit as fixed factor and symptom (yes/no) at each follow-up visit as dependent variable.

### Symptom clusters

The prevalence of symptoms and the overlap between symptom clusters at 3 months follow-up is shown in figure 2. At 3 months, 90.7% of patients reported at least 1 symptom from the physical symptom cluster; this declined significantly to 86.8% at 6 months and to 84.5% at 12 months (p=0.025). Respiratory symptoms were reported by 87.3%, 79.1%, and 76.0% of the patients at 3, 6, and 12 months, respectively (p<0.001). In the fatigue symptom cluster, 68.3% of the patients reported a symptom at 3 months, 67.8% at 6 months, and 67.6% at 12 months (p=0.082). A symptom from the cognitive symptom cluster was reported in 71.8% of the patients at 3 months, 70.0% at 6 months, and 74.2% at 12 months (p=0.452).

At 3 months after hospital discharge, 218 (56.3%) reported symptoms in all 4 symptoms clusters and 292 (75.5%) in 3 clusters. Symptoms in the physical and respiratory symptom clusters most frequently overlapped (figure 3A). The majority of patients with fatigue also experience cognitive symptoms (86.8%), and vice versa (83.4%). Isolated symptoms were rare, but concerned most frequently fatigue in 21 (5.3%) patients or physical symptoms in 18 (4.6%) patients. Females more frequently report symptoms in all four clusters than males (63% versus 52%, p<0.001) (figure 3B and 3C). Fatigue and cognitive symptoms were more frequent in female than in males (80.2% versus 68.6%, p=0.002), and 74.5% versus 68.6%, p=0.009, respectively), and so were the frequency of respiratory (85.9% versus 85.5%, p=0.022) and physical symptoms (90% versus 88.5%, p=0.002).

**Figure 3:**
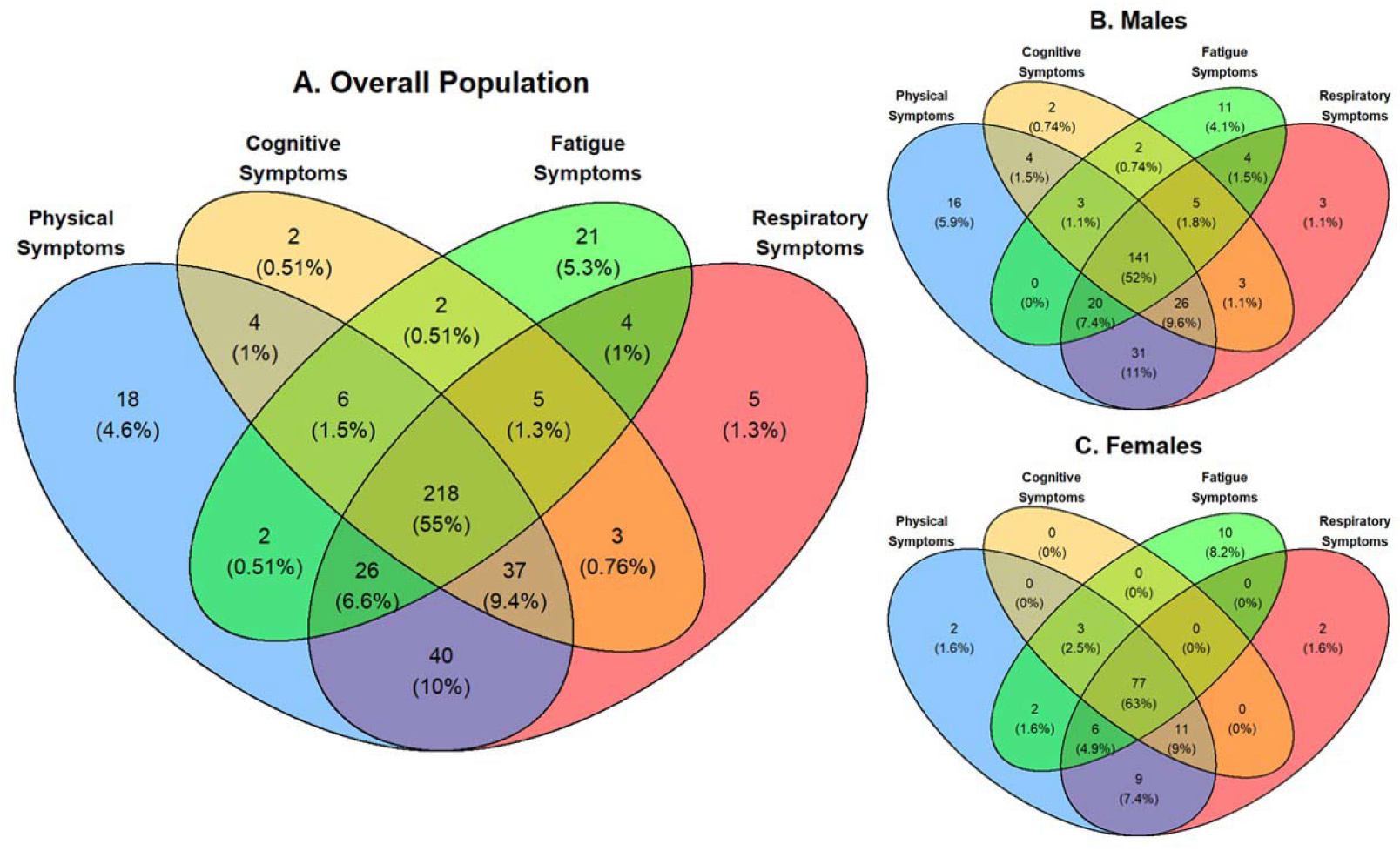
Venn Diagrams showing overlap between the symptom clusters (physical symptoms, cognitive symptoms, fatigue symptoms and respiratory symptoms for the entire cohort (A) and for males (B), and females (C). Data are presented as n (%) indicating the number of patients with symptoms.

In table 3 patient and clinical characteristics of patients hospitalized for COVID-19 at three months follow-up are presented across the number of symptoms clusters affected. The majority of patients 89%) experiences symptoms in 2 or more clusters. Several trends can be noticed in with increasing number of symptoms clusters affected; more in females, patients with non-European background, employment, co-morbidities and with lower CRP, lower D-dimer and more severely affected chest X-ray upon admission. No association seems present with LOS, ICU admission and ventilation and with COVID-19 directed treatment. As numbers per group/characteristic were limited, differences were not statistically assessed and these trends should be considered as explorative and hypotheses generating.

**Table 3.**
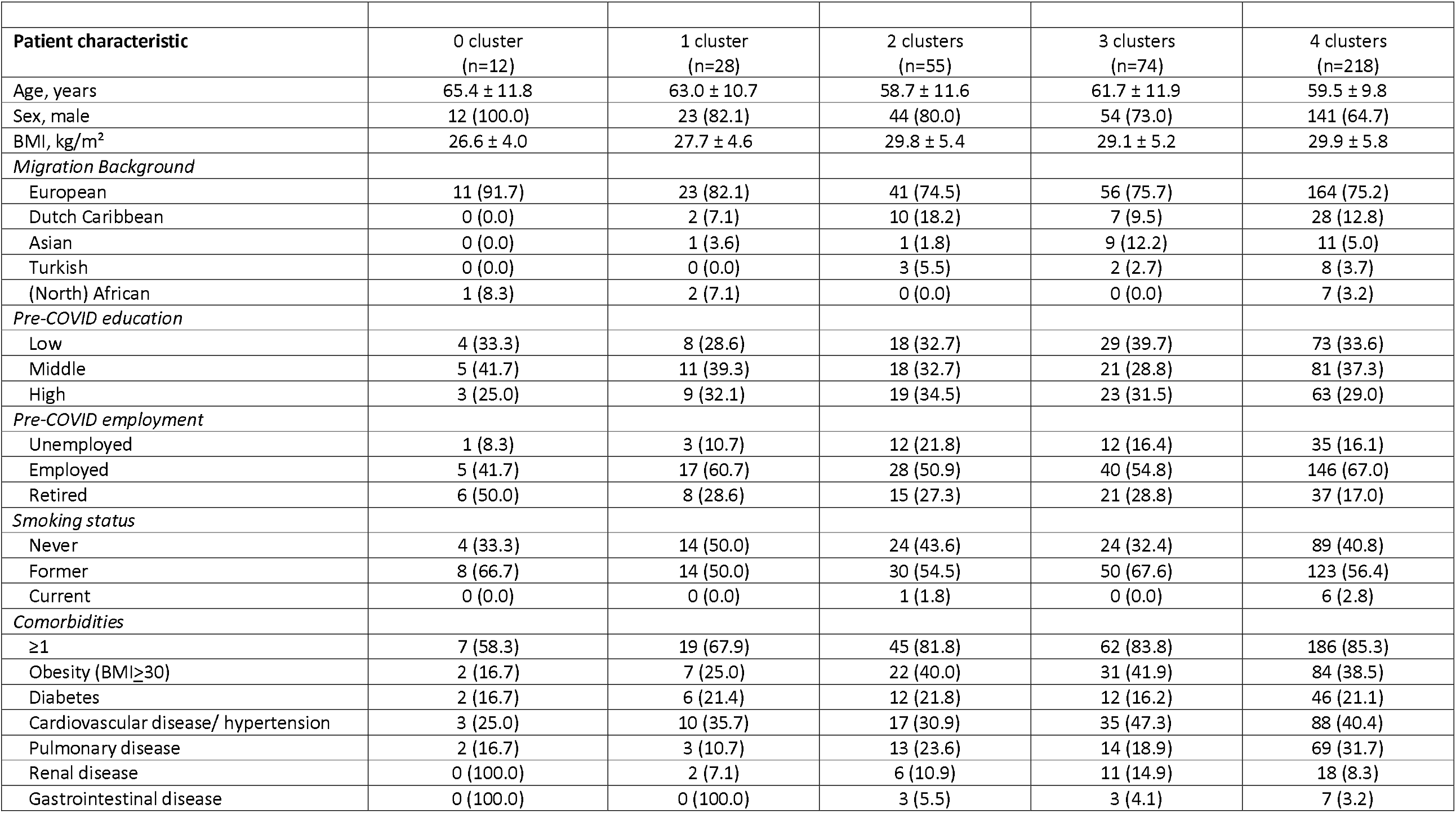
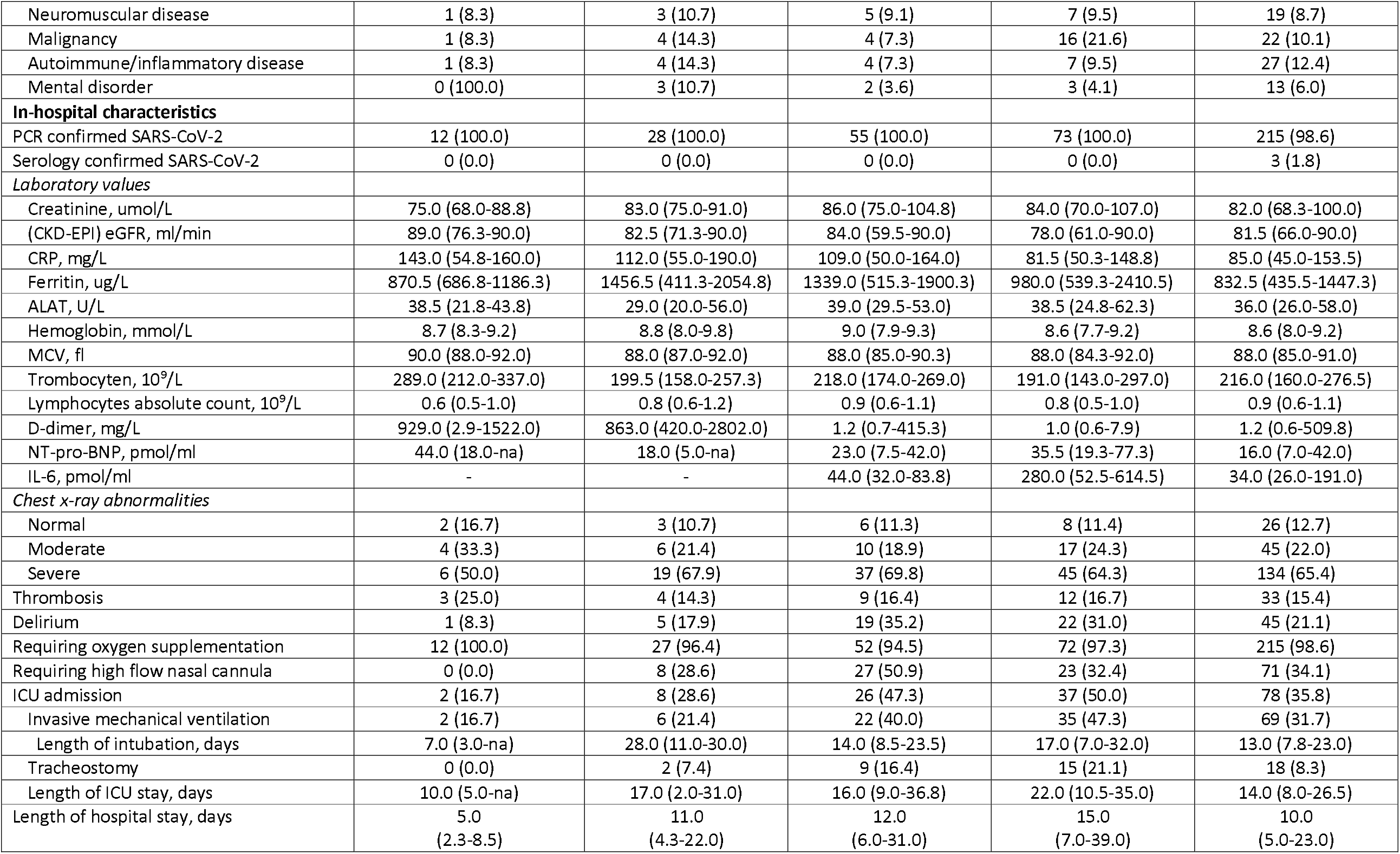
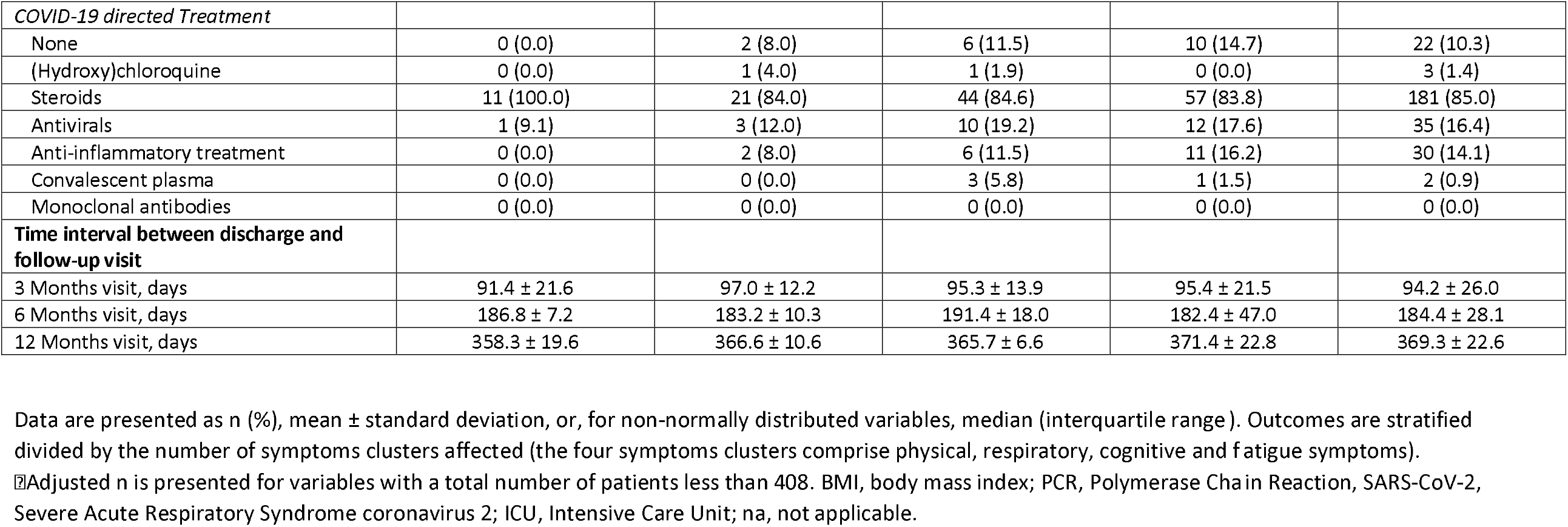
Patient and clinical characteristics of patients hospitalized for COVID-19 divided by the number of symptoms clusters affected.

### Determinants of persisting symptoms

Out of the physical symptom cluster, muscle weakness was the most frequently reported symptom at 3 months after hospital discharge. Patients who were female (OR(95%CI):2.66 (1.62, 4.37), p<0.001), had a longer LOS hospital (1.04 (1.02, 1.05), p<0.001)) were more likely to experience muscle weakness at 3 months after hospital discharge and patients who received steroids as treatment during hospitalization (0.53 (0.29, 0.96), p=0.036) were less likely to experience muscle weakness (figure 4). Out of the respiratory symptom cluster, exertional dyspnea was the most prevalent symptom. We were unable to perform valid multivariable logistic regression on this outcome given the high prevalence of this symptom (81.5%) at 3 months after hospital discharge. Fatigue was the most prevalent symptom in the fatigue symptom cluster. Patients who were female (2.76 (1.61, 4.76), p<0.001) and/or had comorbidities (2.19 (1.24, 3.87), p=0.007) were more likely to develop fatigue symptoms, while patients who were retired (0.38 (0.22, 0.65), p=0.001) were less likely to develop fatigue symptoms at 3 months after hospital discharge (figure 4).

**Figure 4.**
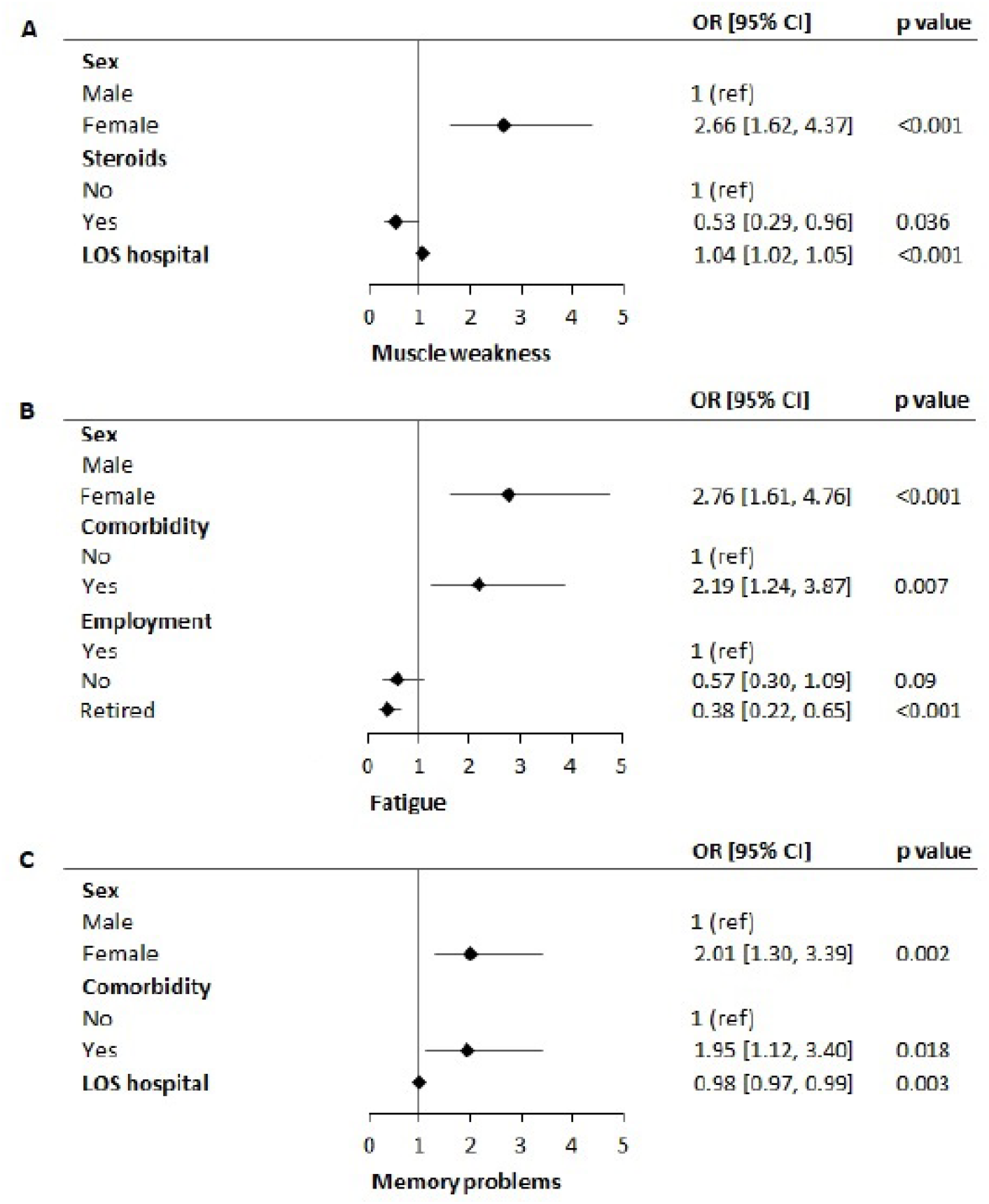
Forest Plot of the patient and admission characteristics associated with the most prevalent symptoms for the physical, fatigue and cognitive symptoms clusters obtained by multivariable logistic regression analyses

At 3 months, memory problems was the most frequently reported symptom in the cognitive symptom cluster. Patients who were female (2.01 (1.30, 3.39), p=0.002), had a shorter length of stay (0.98 (0.97, 0.99), p=0.003), and/or had comorbidities (1.95 (1.12, 3.40), p=0.018) were more likely to experience memory problems at 3 months after hospital discharge (figure 4).

## DISCUSSION

Up to 12 months after hospitalization for COVID-19 over 90% of patients suffer from at least 1 persisting symptom. Muscle weakness, exertional dyspnea, fatigue, and memory and concentration problems were the most prevalent symptoms with reporting rates over 50% of the patients at one of the time points. Although several physical and respiratory symptoms (muscle weakness, hair loss, exertional dyspnea) declined significantly over time, others - including fatigue and cognitive symptoms - persisted.

Our findings support the observation from a recent meta-analysis describing that the short-term prevalence of persisting symptoms were similar to long-term prevalence of symptoms, up to six months after hospital discharge (8). Persisting symptoms are a common feature of COVID-19, especially after hospitalization. To date, long-term data regarding persisting symptoms at 12 months and beyond are limited. In a cohort study from Wuhan, the proportion of persistent symptoms was shown to decrease from 68% at 6 months to 49% at 12 months after hospitalization (10). Although this finding appears to contrast our findings, their cohort contained only 1% of patients that had received mechanical ventilation compared to 35.6% in our cohort. The severity of the acute COVID-19 infection is increasingly recognized to be associated with a larger proportion, and longer duration of persisting symptoms and should thus be taken into account when comparing studies (16, 17). Our study unfortunately shows a much less optimistic picture regarding recovery over time, with high prevalence of persisting symptoms. More and longer term follow-up results will be collected to obtain even more insight into the future outlook of these patients.

Looking into determinants, female gender was the most important predictor of persistent symptoms. Earlier, we have demonstrated a relation of female gender with increased risk for fatigue up to six months after discharge (7). Now we extend these findings to other symptoms and show that females more frequently experience symptoms from multiple symptom clusters 3 months after hospitalization. Previous studies have also demonstrated that female gender was associated with an increased number of persisting symptoms (10, 18). It is frequently stated that, while acute cases of COVID-19 tend to be most severe in older males, PCC seems to be more frequent in younger females. Age however, was not found to be a determinant of persistent symptoms in our cohort, neither in other studies after adjustment for confounders (10, 18). Also, it is necessary to bear in mind that there may be a bias in symptoms reporting between males and females (19). We also found presence of comorbidity to be associated with increased fatigue and memory problems, but this was not found by others (10). It is well possible that pre-existent health problems make patients more vulnerable for unfavorable outcome after severe illness.

Although numbers of acute COVID-19 may eventually decrease with increasing immunity in the population, our findings point out that consequences will be long felt by many. As challenges in vaccination programs worldwide continue to hinder effective control measures, the number of people with PCC will only continue to increase. Vaccination may play an important role not only to prevent new infections, but also in preventing PCC, as it was recently demonstrated that post-vaccination breakthrough infections are less likely to be associated with symptoms persisting for more than 28 days (20).

The best approach to PCC is unclear. As symptoms range from mild to severe and are very diverse in nature, there is no one size fits all treatment possible. Although we grouped symptoms into clusters, there are currently no universal recognized phenotypes, diagnostic criteria, minimal severity scores, or a diagnostic test to establish a diagnosis of PCC. Establishing more objective and evidence-based definitions and phenotypes of PCC will be necessary to compare findings across cohorts and settings, and to establish evidence based interventions. Also, the impact of prior symptoms and prior comorbidity need to be taken into account, just as the expected effects of hospitalization.

There are currently many unknowns regarding PCC, including the underlying mechanisms. Current theories on PCC include virus-specific pathophysiologic changes, immunologic aberrations and inflammatory damage in response to the acute infection, and expected sequelae of post-critical illness (21). Indeed, it is very hard to differentiate between the expected sequelae, such as described in the post-intensive care syndrome, and the sequelae that are specific for COVID-19 (22). Nonetheless, in a large analysis on electronic health records data, key features of long-COVID (e.g. breathlessness and fatigue) were more frequently reported after COVID-19 infection than in matched controls after Influenza infection (13). Overall, it is becoming increasingly clear that the sequelae after COVID-19 are more prevalent than after most other types of infections, persist for a long time, and have a major impact on the burden of disease and health care.

Our study has several strengths and limitations. We followed a large cohort of patients in a longitudinal design at 3, 6, and 12 months after hospital discharge. Currently, long-term follow-up data are scarce. As the study is still ongoing, data were not complete for the entire cohort; also, the initial patients were generally recruited between 3 and 6 months after hospital discharge, resulting in unequal groups at different time points. We therefore included only participants with data of at least two study measurements, and used GEE models to make maximal use of all data and to investigate how symptoms developed over time.

One inclusion criterium was that patients or their relatives had to be able to communicate in Dutch or English. Therefore, there is underrepresentation of individuals with a migration background in this study compared to the community where this cohort was established. Nonetheless, 24% of the participants in this cohort had a migration background; migration background was not a predictor of residual symptoms in this study. Another limitation is that our results are only generalizable to hospitalized patients. Also, we did not have patient scores on the severity of complaints. Even though symptoms may persist for considerable time, the severity may very well decrease over time, as was also shown in other studies (16).

To summarize, a large number of post COVID-19 patients experienced persistent symptoms up to 12 months after hospitalization for COVID-19. Whereas physical and respiratory symptoms show slow gradual decline, fatigue and cognitive symptoms did not evidently resolve over time. This finding stresses the importance of finding the underlying causes and effective treatments for PCC on the one hand, and adequate COVID-19 prevention on the other hand. Large and long-term cohort studies together with broader consortia are urgently needed to help better understand the trajectory, complications, and biological mechanisms that drive persistent symptoms after COVID-19 infection.

## Data Availability

All data produced in the present study are available upon reasonable request to the authors

## Appendix 1

### Corona Symptom Checklist

The following complaints may be experienced after COVID-19 infection. Check for each question whether you experienced the complaint at this moment <u>**and**</u> is new since the COVID-19 infection. When you experienced the complaints already before the infection and this did not change, please check “no”. When the complaints worsened or is new after infection, please check “yes”.

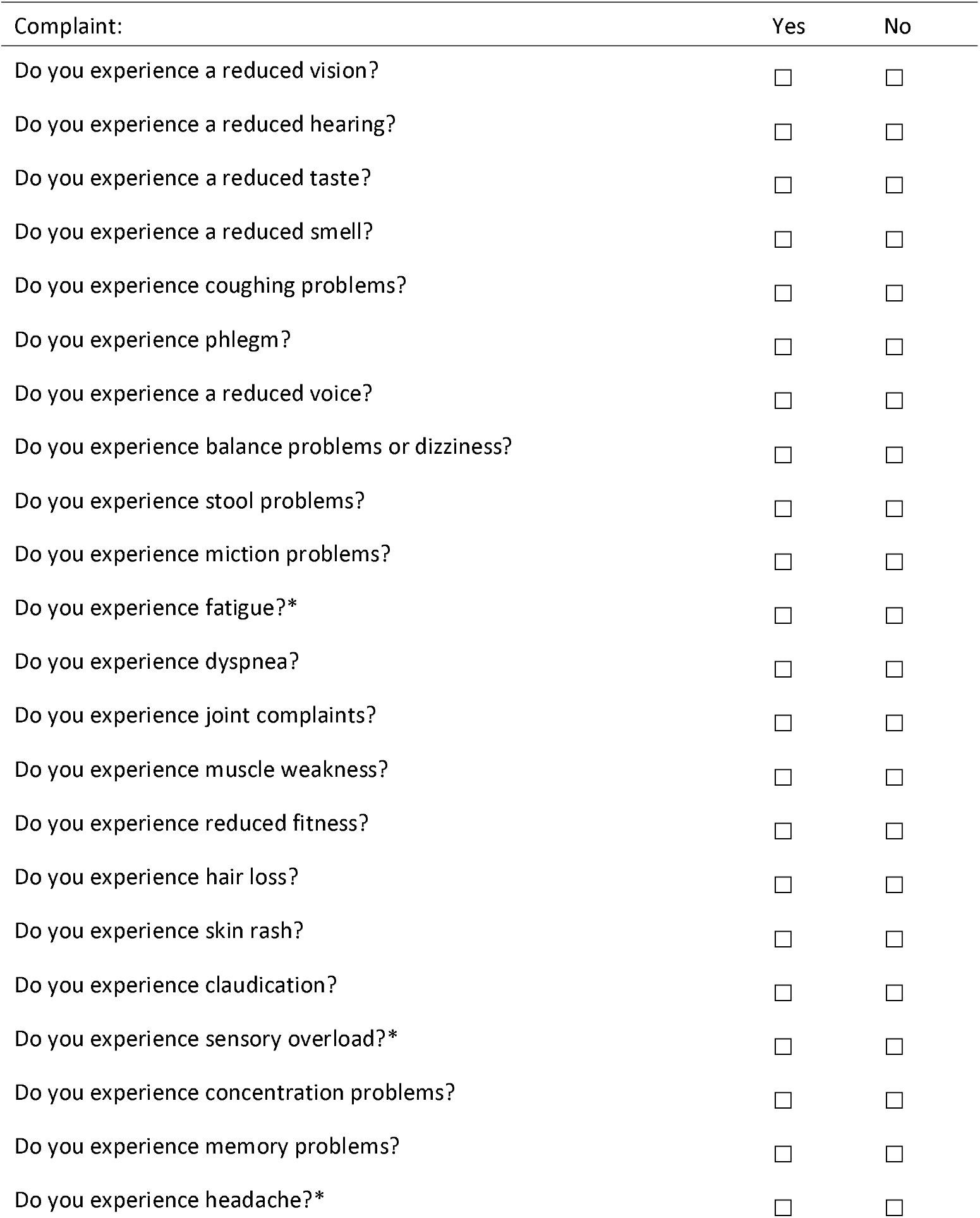
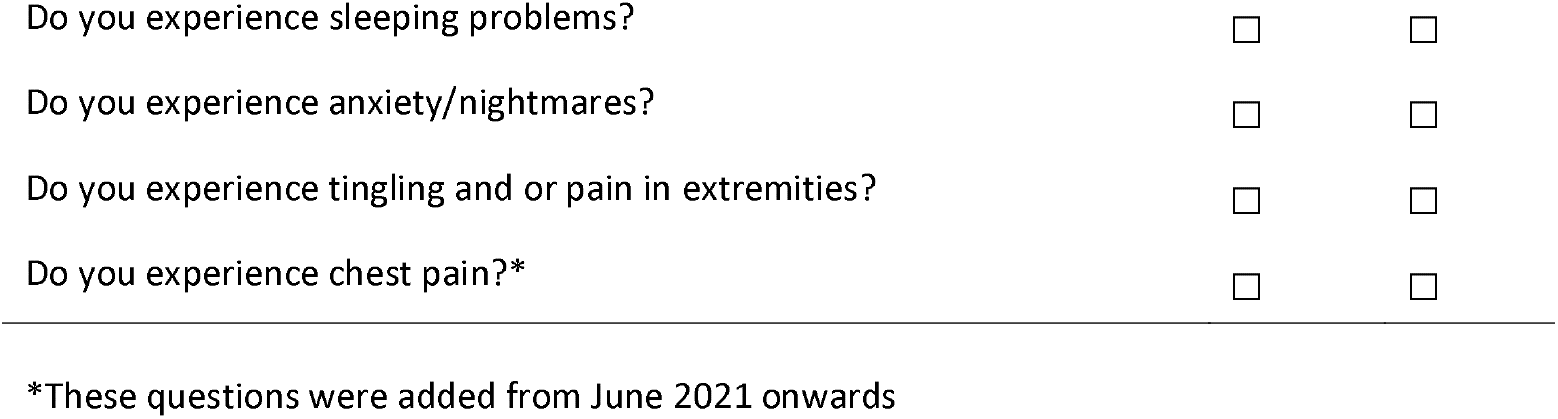

| Complaint: | Yes | No |
| --- | --- | --- |
| Do you experience a reduced vision? | ☐ | ☐ |
| Do you experience a reduced hearing? | ☐ | ☐ |
| Do you experience a reduced taste? | ☐ | ☐ |
| Do you experience a reduced smell? | ☐ | ☐ |
| Do you experience coughing problems? | ☐ | ☐ |
| Do you experience phlegm? | ☐ | ☐ |
| Do you experience a reduced voice? | ☐ | ☐ |
| Do you experience balance problems or dizziness? | ☐ | ☐ |
| Do you experience stool problems? | ☐ | ☐ |
| Do you experience miction problems? | ☐ | ☐ |
| Do you experience fatigue?* | ☐ | ☐ |
| Do you experience dyspnea? | ☐ | ☐ |
| Do you experience joint complaints? | ☐ | ☐ |
| Do you experience muscle weakness? | ☐ | ☐ |
| Do you experience reduced fitness? | ☐ | ☐ |
| Do you experience hair loss? | ☐ | ☐ |
| Do you experience skin rash? | ☐ | ☐ |
| Do you experience claudication? | ☐ | ☐ |
| Do you experience sensory overload?* | ☐ | ☐ |
| Do you experience concentration problems? | ☐ | ☐ |
| Do you experience memory problems? | ☐ | ☐ |
| Do you experience headache?* | ☐ | ☐ |
| Do you experience sleeping problems? | ☐ | ☐ |
| Do you experience anxiety/nightmares? | ☐ | ☐ |
| Do you experience tingling and or pain in extremities? | ☐ | ☐ |
| Do you experience chest pain?* | ☐ | ☐ |

\*These questions were added from June 2021 onwards

## Acknowledgements

We would like to extend our gratitude to all the participants of the ongoing CO-FLOW study for their efforts, as well as all the entire group of measurement-assistants that currently assists in CO-FLOW: G.W.M. Broeren, R.M.B. Imkamp, J. Andela, L. Bierman, T. Huijboom, A. Luijckx, S. Roovers, I. Simons, and L. van Veggel

## Funding

The study is funded by the COVID-19 Program Care and Prevention of The Netherlands Organization for Health Research and Development (ZonMw, grant number 10430022010026), and Rijndam Rehabilitation and Laurens (both in Rotterdam, The Netherlands). The review committee COVID-19 of the funding body has independently reviewed the study protocol. None of the funders have a role in the study design, nor are involved in analysis and interpretation of data or writing of the manuscript.

## Author Contributions

LB, JB, MHK, HB, MH devised the main conceptual ideas and proof outline for this manuscript; LB, JB, MH included the patients; LB, JB contributed to data collection and aggregation; MH and JV designed the figures; LB, JB, MHK performed the statistical analyses; LB, JB, MHK, HB, MH wrote the manuscript. All authors were involved in the main study design, discussed the results, commented on the manuscript and approved the final manuscript.

## Authors of the CO-FLOW collaboration Group

### CO-FLOW collaboration Group

Michel E. van Genderen^1^ Jasper van Bommel^1^, Diederik A.M.P.J. Gommers^1^, Erwin Ista^2,3^, Robert van der Stoep^4^, Rutger Osterthun^5,6^, Markus P.J.M. Wijffels^6^, Marieke M. Visser^6^, Janette J. Tazmi-Staal^7^, Eva G. Willems^7^, Roxane Heller^8^, Shai A. Gajadin^9^, Wouter J.B. Blox^10^, Laurien Oswald^11^, Sieshem Bindraban^11^, Rob Slingerland^12^, Herbert J. van de Sande^13^, Ronald N. van Rossem^14^, Stephanie van Loon-Kooij^14^, L. Martine Bek^5^, Julia C. Berentschot^15^, Merel E. Hellemons^15^, Susanne M. Huijts^15^, Joachim G.J.V. Aerts^15^, Majanka H. Heijenbrok-Kal^5,6^, Rita J.G. van den Berg-Emons^5^, Gerard M. Ribbers^5,6^

### ORCID IDs

<u>0000-0001-5668-3435</u> Michel E. van Genderen

<u>0000-0001-8408-0500</u> Jasper van Bommel

<u>0000-0001-6808-7702</u> Diederik A.M.P.J. Gommers

<u>0000-0003-1257-3108</u> Erwin Ista

<u>0000-0001-9809-7311</u> Rutger Osterthun

<u>0000-0003-1472-7158</u> Eva G. Willems

<u>0000-0003-2706-0639</u> Laurien Oswald

<u>0000-0001-8257-7070</u> L. Martine Bek

<u>0000-0002-6666-5027</u> Merel E. Hellemons

<u>0000-0002-6652-6254</u> Susanne M. Huijts

<u>0000-0001-6662-2951</u> Joachim G.J.V. Aerts

<u>0000-0002-2982-4404</u> Majanka H. Heijenbrok-Kal

<u>0000-0001-6433-3398</u> Rita J.G. van den Berg-Emons

<u>0000-0002-6114-349X</u> Gerard M. Ribbers

### Author details

^1^Department of Adult Intensive Care Medicine, Erasmus MC, University Medical Center Rotterdam, The Netherlands.

^2^Departments of Pediatrics and Pediatric Surgery, Intensive Care Unit, Erasmus MC Sophia Children’s Hospital Rotterdam, The Netherlands.

^3^Department of Internal Medicine, section Nursing Science, Erasmus MC, Erasmus University Medical Center Rotterdam, The Netherlands.

^4^Department of Physical Therapy, Erasmus MC, University Medical Center Rotterdam, The Netherlands.

^5^Department of Rehabilitation Medicine, Erasmus MC, University Medical Center Rotterdam, The Netherlands.

^6^Rijndam Rehabilitation, Rotterdam, The Netherlands.

^7^ Laurens Intermezzo, Rotterdam, The Netherlands.

^8^Department of Respiratory Medicine, Ikazia Hospital, Rotterdam, The Netherlands.

^9^Department of Respiratory Medicine, IJsselland Hospital, Capelle aan de IJssel, The Netherlands.

^10^Department of Respiratory Medicine, Albert Schweitzer Hospital, Dordrecht, The Netherlands.

^11^Department of Respiratory Medicine, Franciscus Gasthuis & Vlietland, Rotterdam, The Netherlands.

^12^Department of Respiratory Medicine, Maasstad Hosptial, Rotterdam, The Netherlands.

^13^Aafje Nursing Home, Rotterdam, The Netherlands.

^14^Department of Respiratory Medicine, Reinier de Graaf Gasthuis, Delft, The Netherlands.

^15^Department of Respiratory Medicine, Erasmus MC, University Medical Center Rotterdam, The Netherlands.

## Notes

### Competing Interest Statement

Dr. Aerts reports personal fees and non-financial support from msd, personal fees from bms, personal fees from boehringer ingelheim, personal fees from amphera, personal fees from eli-lilly, personal fees from takeda, personal fees from bayer, personal fees from roche, personal fees from astra zeneca, outside the submitted work; In addition, Dr. Aerts has a patent allogenic tumor cell lysate licensed to amphera, a patent combination immunotherapy in cancer pending, and a patent biomarker for immunotherapy pending.
Other authors have nothing to disclose.

### Clinical Protocols

https://pubmed.ncbi.nlm.nih.gov/34419032/

### Author Declarations

The Medical Ethics Committee of the Erasmus Medical Center (MC) approved this study (MEC-2020-0487).

